# Changes in the quality of cancer care as assessed through performance indicators during the first wave of the COVID-19 pandemic in 2020: a Scoping Review

**DOI:** 10.1101/2022.02.23.22271303

**Authors:** Ana Sofia Carvalho, Óscar Brito Fernandes, Mats de Lange, Hester Lingsma, Niek Klazinga, Dionne Kringos

## Abstract

**Objectives:** Summarize performance indicators used in the literature to evaluate the impact of the COVID-19 pandemic on cancer care (January-June 2020), and to assess changes in the quality of care as assessed via selected indicators.

**Methods:** Scoping review. Indicators and their reported trends were collated following the cancer care pathway.

**Results:** Database searches retrieved 6277 articles, 838 articles met the inclusion criteria, and 135 articles were included after full-text screening, from which 917 indicators were retrieved. Indicators assessing the diagnostic process showed a decreasing trend: from 33 indicators reporting on screening, 30 (91%) signalled a decrease during the pandemic (n=30 indicators, 91%). A reduction was also observed in the number of diagnostic procedures (n=64, 58%) and in the diagnoses (n=130, 89%). The proportion of diagnoses in the emergency setting and waiting times showed an increasing trend (n=8, 89% and n=14, 56%, respectively). Nine indicators (64%) showed stability in cancer stages distribution. A decreasing trend in the proportion of earliest stage cancers was reported by 63% of indicators (n=9), and 70% (n=43) of indicators showed an increasing trend in the proportion of advanced-stage cancers. Indicators reflecting the treatment process signalled a reduction in the number of procedures: 79% (n=82) of indicators concerning surgeries, 72% (n=41) of indicators assessing trends in radiotherapy, and 93% (n=40) of indicators related to systemic therapies. Modifications in cancer treatment were frequently reported: 64% (n=195) of indicators revealed changes in treatment. Ten indicators (83%) signalled a decreasing number of hospital admissions.

**Conclusion:** Health systems struggled to ensure continuity of cancer care. As this pandemic keeps evolving, the trends reported over the first 6 months of 2020 provide an argument to monitor these changes closely. This information needs to be transparent, standardised, and timely, allowing to monitor quality and outcomes of care during crises and inform policy responses.

## Introduction

The COVID-19 outbreak was declared a public health emergency of international concern by the World Health Organization on the 30^th^ of January, 2020 [1]. Since the beginning of the pandemic, global health systems struggled to cope with the high numbers of patients infected with SARS-CoV-2, while maintaining adequate care for non-COVID patients [2].

Cancer comprises a high burden to populations and health systems, being the second cause of death globally [3], and in Organisation for Economic Co-operation and Development (OECD) countries [4]. The COVID-19 pandemic has widely affected cancer care delivery. Substantial declines in the number of cancer diagnoses have been reported in the Netherlands [5, 6], Spain [7], Belgium [8], and Denmark [9]. While trying to minimize the risk of COVID-19 disease in cancer patients, changes in practice were pursued by oncologists, according to each setting’s capacity and recommendations released by oncology societies [10–12]. Changes in the treatment prescribed were reported, by delaying, changing treatment modality, duration, or mode of delivery [13].

Improving cancer care was already on the international health agenda prior to this pandemic, namely in the 2030 Agenda for Sustainable Development adopted in 2015 at the United Nations Sustainable Development Summit [14], in the Resolution “Cancer prevention and control in the context of an integrated approach” approved in 2017 by the World Health Assembly [15]. At the same time, systematic efforts to capture the outcomes of cancer care globally via cancer registries are in place such as the CONCORD study [16, 17]. The OECD reports on cancer care as part of its *Health at a Glance* Series [4] and European Union (joint) actions have taken place [18]. During the COVID-19 pandemic, efforts continued to be pursued to improve prevention, diagnostics, treatment, and the quality of life of cancer patients, among others with the launch of the “Europe’s Beating Cancer Plan” in November 2020 [19] and initiatives to monitor and report upon inequalities, such as the European Cancer Inequalities Registry [20] and the launch of the European Commission’s Knowledge Centre on Cancer [21]. These efforts are necessary to tackle the cancer crisis and to address the backlog this pandemic is creating [22–24].

One key area that needs strengthening is the development of performance indicators, its standardization, and timely collection by health systems data infrastructures [25], to efficiently inform on changes in the quality of care provided, allowing for comparisons within and between countries’ health systems. Although improving outcomes like 5-year survival is the ultimate aim, for guiding health care delivery systems towards that goal, process and cancer-pathway based information is essential.

The literature published on the consequences of the pandemic on cancer care is vast, therefore assessing its impact requires a structured approach. This scoping review is part of a larger project, which focuses on performance indicators on the care for non-communicable diseases (NCD). This study aims to provide a systematic summary of cancer care performance indicators used in the literature, regarding various cancers, across the care pathway, from early detection to outcomes, and with an international coverage. Additionally, we set out to assess the main trends of the changes in the quality of cancer care during the first wave of the COVID-19 pandemic, from January to June 2020, by collecting and collating the performance indicators’ trends across the cancer care pathway.

## Methods

We conducted a scoping review, following Arksey and O’Malley methodological framework [26], further developed by Levac *et al* [27]. Given the lack of standardized methods across countries for data collection on health care system performance assessment and their translation to indicators [25, 28], a scoping review methodology allows to map the large sums and heterogeneity of literature available [27,29,30] and to identify key concepts present in the literature [30], namely cancer care performance indicators. It also enables the reporting of emerging evidence [29], to summarize, and communicate findings [26], including trends revealed by indicators. The PRISMA extension for scoping reviews [31] was used for reporting (S1 Table).

### Eligibility criteria of studies

We considered the following inclusion criteria: 1) studies using empirical data on the use of health services in OECD countries, 2) studies that described health outcomes and/or performance indicators related with NCD during the COVID-19 pandemic, 3) original journal articles using quantitative or qualitative methods (cohort studies, case-control, cross-sectional, case reports, systematic reviews, surveys, and meta-analyses). Studies were excluded if they did not provide empirical data on health services and NCD, namely: 1) editorials and commentaries, 2) prediction models, 3) clinical case reports; 4) diseases management or health services organization guidelines, 5) studies about the impact on healthcare workers, patients diagnosed with COVID-19, children, or pregnant women; 6) studies primarily performed in non-OECD countries. No limitations were set regarding language or year of publication.

### Information sources

MEDLINE and Embase databases were selected to search for this study, given the large number of articles and their comprehensive coverage of the literature on health services delivery. Pilot searches were conducted to identify a list of suitable search terms. An experienced medical information specialist was consulted to improve the search strategy, which was refined with discussion among co-authors. The comprehensive search included search terms grouped by key concepts (COVID-19, pandemic, non-communicable disease, chronic disease, performance indicator, healthcare quality, healthcare utilization, healthcare delivery and other closely related terms). The search was adapted to both databases and conducted by the information specialist on 17-03-2021. The full search strategy for Embase can be found in S2 Doc. Duplicates were removed using EndNote software. Additional articles of relevance were added by hand-searching the reference lists of the included studies.

### Selection of sources of evidence

Titles and abstracts screening was performed independently by two researchers (ASC, OBF) using Rayyan [32]. Studies considered relevant were exported to a spreadsheet to support full-text screening. For this study, only articles related to cancer care were analysed. Full-text screening was performed independently by two researchers (ASC, MdL). The reason for the exclusion of articles was recorded at this point, and both researchers agreed on the excluded studies. In case of doubt, the other co-authors were consulted.

### Data extraction

Data extracted was collated in a spreadsheet (S3 Table) piloted on 15 studies. Prior to extracting data from all studies, two researchers (ASC, MdL) compared data collected from 10 selected articles with the objective of enhancing data extraction consistency among researchers. Then, data was charted independently by ASC and MdL. Extracted data included information on generic and methodological aspects of the article (e.g., authors, title, setting) and information about the indicators collected (e.g., indicator title, and data inclusion/exclusion considerations). For every indicator, we identified the trend reported in the articles (increase/decrease/stable), if any.

### Synthesis of results

We grouped and categorised the indicators following clinical reasoning. These categories were organized according to the different phases of the cancer care pathway: early detection, diagnosis, treatment, and outcomes. Quantitative indicators’ trends were collated, and the percentage of indicators reporting each trend (increase/decrease/stable) was computed for each category. This evidence is presented in the text of this study and in a diagram informing about each phase of the cancer care pathway. Qualitative information extracted from surveys is presented in the text, and it was not considered to compute trends. In the category “changes in treatment” quantitative indicators and qualitative information were grouped and categorized according to clinical reasoning to present an overview of the modifications in cancer treatment reported.

## Results

Database searches retrieved 6277 articles. Of these articles, 838 met the inclusion criteria, from which 197 articles on cancer care were identified. Eight records were identified via hand-searching. After full-text screening, 135 articles were included in this study (Fig. 1).

**Fig. 1.**
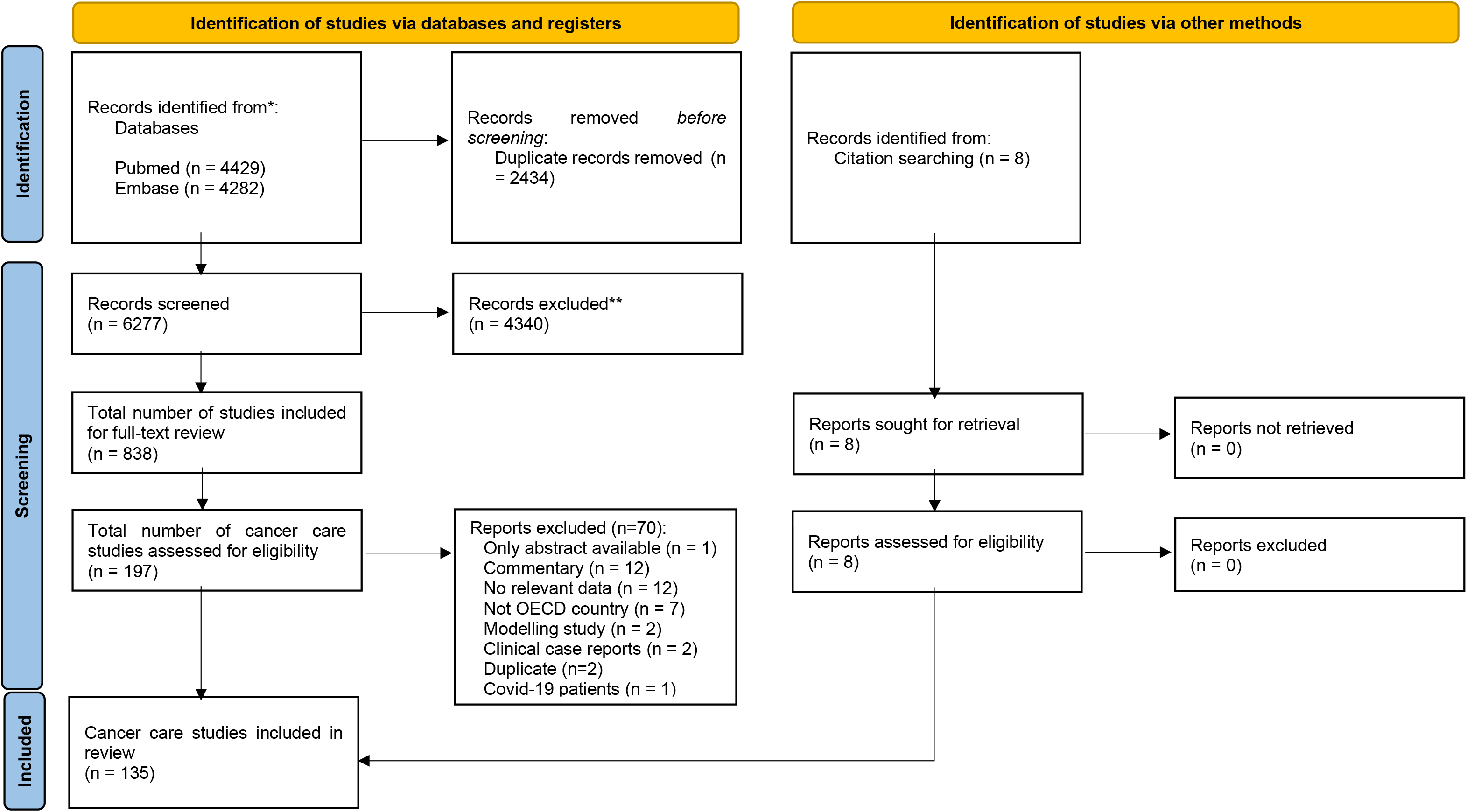
PRISMA flow diagram of the literature search (17-03-2021)

### General characteristics of the included articles

In total, 135 studies were included, reporting on 94 countries (S4 Table). Of these studies, 26 (19%) provided information on multiple countries, from which 14 (10%) specified all the countries included. Of those, 8 (6%) included non-OECD countries. Most of the studies including more than one country were surveys (n=23, 89%). The most frequent countries reported on were Italy (n=36, 29%), US (n=32, 26%) and UK (n=27, 22%) (Fig. 2). Most articles used a retrospective cohort design (n=82, 61%), followed by surveys (n=44, 33%). Surveys were often directed to health professionals (n=37/44, 84%) and to patients (n=6/44, 14%) to a lesser extent. Other study designs that were applied included prospective cohorts (n=5), observational retrospective cohorts (n=3), and a combination of prospective and retrospective cohort (n=1). Studies reported indicators’ trends during the first phase of the COVID-19 pandemic (from January to June 2020) and, in some cases, after the (in many countries implemented) lockdown period (from May to October 2020). The magnitude of each indicator in 2020 was compared with its magnitude in the same period of 2019 (n=51, 38%), to a period immediately before (n=33, 24%) or to the average of the same period in previous years (ranging from 2017 to 2019) (n=26, 19%).

**Fig. 2.**
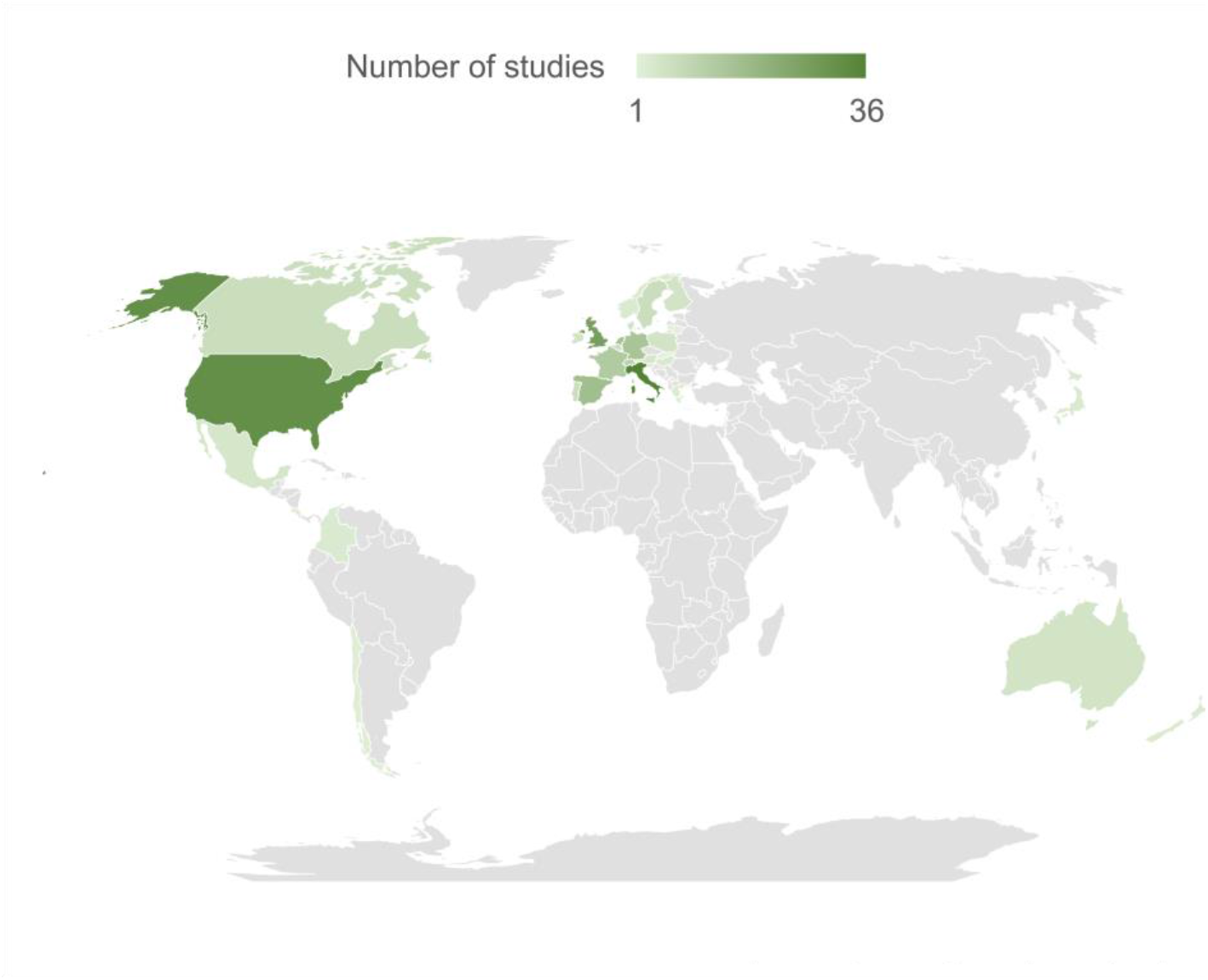
OECD countries reported on, color-graded according to the number of papers (n=122 articles; 90% of included articles).

### Cancer Care Indicators

A total of 917 quantitative indicators from 91 articles were retrieved and grouped into categories (Table 1).

**Table 1.**
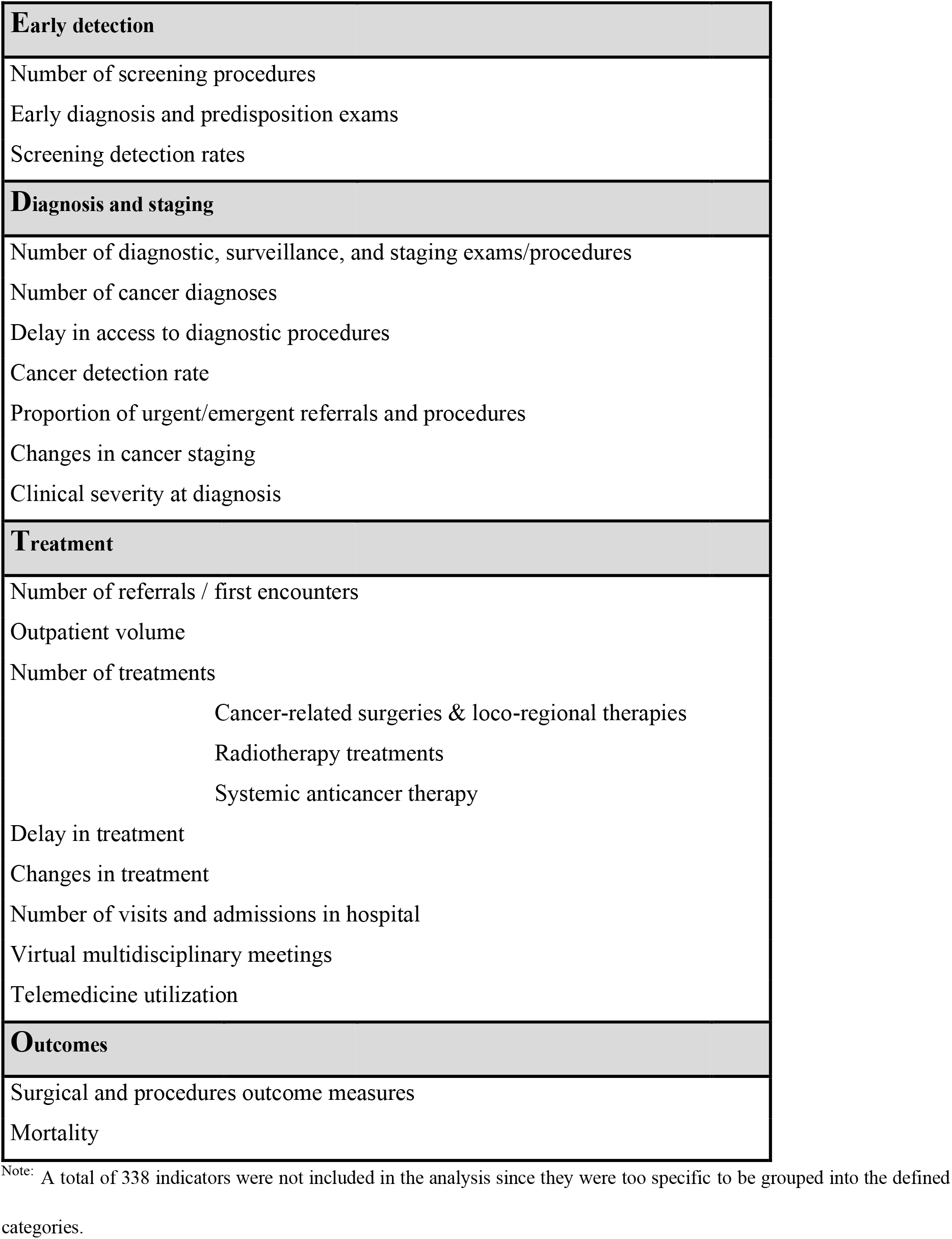
Cancer care indicators’ categories retrieved in the literature review

The first stage of the care pathway looks at **early detection**. Regarding the “number of screening and other early diagnosis procedures”, a total of 33 indicators from 13 articles [33–45] have been reported. Most indicators on the number of screening procedures signaled a decreasing trend (n=30, 91%), namely in US, UK, and Italy. One international survey answered by physicians [46] (including Italy, Iran, Spain, UK, US, China, Denmark, Sweden, and Switzerland) reported the suspension of breast screening programs in all countries, except 2. Only one American paper [38] addressed cervical cancer screening, showing a decrease in number of screenings during stay-at-home order, compared with 2019. Three articles [35,39,42] addressed colorectal cancer screening, from US, UK and Spain and have also revealed a reduction when compared with previous year.

Seven papers [33,34,36–38,40,42] reported on “early diagnosis and predisposition exams”, namely: screening visit for prostate cancer (US), gBRCA testing (Italy), HPV tests rate, low-dose computed tomography and prostate-specific antigen measurement (all from the US). All these indicators have shown a decreasing trend in the number of diagnostic procedures.

“Screening detection rates” were reported by 14 indicators from 5 articles [22,36,37,47,48], from the US and Italy. Most of the indicators (n=9, 75%) showed an increase in screening detection rates. These included an increase in high-risk adenomas and colorectal cancer detection rates during the lockdown period, along with a decrease in low-risk adenoma detection rates in one Italian study [47].

The next stage of the care pathway focuses on **diagnosis and staging.** The “number of diagnostic, surveillance and staging procedures” was reported in 17 articles [41,43–45,48–60] (from the UK, Italy, US, France, Australia, the Netherlands, Turkey, Ireland, and Slovenia), comprising a total of 90 indicators. Most of those indicators (n=58, 64%) showed a decreasing trend in the number of procedures, namely cystoscopy, diagnostic mammograms, breast cancer wire-guided biopsy, gastroscopy, colonoscopy, computed tomography (CT), magnetic resonance imaging (MRI), and endoscopic retrograde cholangiopancreatography.

A total of 147 indicators from 35 articles [9,35,39,47,48,52,54,55,59–83] reported on the “number of cancer diagnosis”. Most of these indicators (n=130, 89%) signalled a decrease in the number of cancer diagnoses. Data came from registries in the Netherlands, Denmark, Germany, and Italy, cytology laboratories, tumor boards numbers, and administrative sources. One international survey to laboratories from 23 countries [66] showed an absolute reduction in the number of cytological samples regarding all anatomic sites.

Five studies [37,55,67,75,82] (from the US, UK, France, and Italy) addressed the number of cancer diagnoses after the lockdown period (10 indicators). Three of these indicators (30%) showed a higher number of diagnoses, when compared with the period of lockdown.

Five surveys [84–88] reported “delays concerning different aspects of cancer diagnostics”. One international survey [85] focusing on colorectal cancer care, with professionals from 84 countries, reported delays in radiologic exams and endoscopic procedures. Other surveys mentioned limited access to hospital facilities (Italy) [89], delays in tissue diagnosis (UK) [88], delays in diagnostics of patients with neuroendocrine tumors (Germany, Austria, Switzerland) [87], and genetic testing or counseling (US) [86]. The other two articles [43, 90] addressed the delays in access to diagnostics, comprising a total of 9 quantitative indicators, from which 8 (89%) signalled an increase in the waiting time to diagnostic procedures.

The “cancer detection rate in referrals and diagnostic exams” was assessed in 5 articles, from UK [59, 91], Italy [48], Ireland [43], and one international survey of 23 laboratories worldwide [66]. The 13 indicators collected signalled an increasing trend in cancer detection rate in 9 indicators (69%). The survey reported an increase of 5.5% in the malignancy rate in nongynecological samples, when compared with the corresponding period in 2019.

Twenty-five indicators from 5 studies [39,57,59,92,93] (from Spain, UK, Australia, and Croatia) reported on the proportion of “urgent/emergent referrals and procedures”. Most of these (n=14, 56%) showed an increase in the proportion of urgent procedures (endoscopy and colonoscopies), diagnosis in emergency setting or operations that followed an emergency admission.

The indicators that reported on “changes in cancer staging” were grouped in 3 different categories: general staging indicators (14 indicators from 8 articles [75,77,81,94–98]), proportion of earliest-stage cancers (35 indicators from 9 articles [41,71,81,99–104]), and proportion of advanced-stage cancers (61 indicators from 17 articles [39,57,63,66,77,79,87,92,93,95–102]).

Most of the general staging indicators (n=9, 64%) showed stability in cancer stages distribution at diagnosis, when comparing the pre-and post-lockdown periods (data from Italy, US, UK, France, US, and Portugal). These indicators included stages of gynecological cancer, breast cancer, lung cancer, and hepatocellular carcinoma. With respect to the proportion of earliest stage cancers, most of the indicators (n=9, 63%) showed a lower proportion of these cancers when compared with pre-pandemic period, 9 (26%) signalled a stable trend, and 4 (11%) showed an increasing trend. From the indicators reporting on the proportion of advanced-stage cancers, 43 (70%) showed an increasing trend in this proportion after the beginning of the pandemic, 14 (23%) signalled a stable proportion, and 4 (7%) reported a decreasing trend.

Fourteen indicators from 7 articles [41,47,57,69,97,100,105] evaluated the “clinical severity at diagnosis”, which included symptoms, scores, and biomarkers. The majority (n=8 indicators, 57%) showed patients presenting in a more severe clinical condition than before the pandemic, namely in US (endometrial cancer), Portugal (hepatocellular carcinoma), Italy, and Turkey (colorectal cancer).

The following stage of the care pathway reported on is **cancer treatment**. A total of 41 indicators (from 9 articles [35,60,65,72,91,96,103,106,107]) reported on the “number of cancer patients’ referrals”. Most of the indicators (37, 90%) signalled a decrease in the number of first encounters for oncological examination, namely in Slovenia, UK, US, France, Spain, and the Netherlands. Four surveys (from UK [88], US, Italy [108] and one international study [109] reported a decrease in the number of new referrals.

A total of 47 indicators were identified from 14 articles [35,42,44,52,55,60,65,90,91,93,96,106,110,111] regarding the outpatient care of patients diagnosed with cancer. Of these indicators, 46 (93%) showed a decrease in the number of outpatient visits (in Korea, US, France, UK, Spain, Slovenia, and Italy). Thirteen surveys [13,46,70,86,87,89,108,112– 117] disclosed information concerning outpatient care. Ten were answered by oncologists and 3 were answered by patients. The latter studies reported consequences on treatment or follow-up (Netherlands) [114], namely treatment adjustment, postponement, delay, or discontinuation; delay in routine or follow-up clinic appointment (US) [86] and postponements of physician appointments (Germany). A substantial percentage of physicians reported cancellation or deferral of follow-up visits.

Regarding the “volume of cancer treatment”, indicators regarding the 3 main therapeutic components were reported: surgeries and loco-regional therapies, radiotherapy, and systemic therapy.

Concerning surgeries and loco-regional therapies, 106 indicators were identified from 30 articles [42,48–51,55,62,65,65,75,81,90,92–94,97–99,101–103,111,118–125]. Of those, 82 indicators (79%) showed a reduction in the number of treatment procedures, namely in Italy, France, Germany, Ireland, Netherlands, Spain, Turkey, UK, US, Australia, and in 1 international study. Nineteen articles were surveys directed to physicians [46,58,70,78,84,85,88,90,108,115,126–134], out of which 8 were international. All have shown significant reductions in surgical activity regarding different cancers.

Regarding radiotherapy treatments, 57 indicators from 8 articles [55,90,92,121,135–138] were identified. Most of these indicators signalled a reduction of the number of treatments (n=41, 72%), namely in UK, US, and Canada. Five international surveys to physicians [112,114,126,132,137] also reported that this type of cancer therapy was affected.

With respect to systemic anticancer therapy, 43 indicators were identified from 9 articles [42,53,55,72,96,106,111,139,140], concerning Italy, France, UK, Spain, and the US. The majority (n=40, 93%) showed a decrease in requests for initial treatment and in the number of chemotherapy administrations. Six surveys [58,109,112,115,134,141] reported on this treatment modality. One study including 54 countries [46] showed that 88.2% of centers reported a reduction in their usual level of care, while 9.83% of those reported lack of access to medications. A European study with 29 countries [109] reported that 6% of centers revealed shortages of drugs.

A total of 28 indicators (from 3 articles) [55,139,140] reported on systemic anticancer therapy after lockdown ending (from May to October 2020). Of these indicators, 14 (50%) reported an increase in the number of treatments (in UK and France).

Regarding “delay in treatment”, a total of 42 indicators from 18 articles [39,52,72,77,81,89,91,92,96,97,99,101,138,142–146] were identified. Of those, 18 (43%) reported an increasing trend in waiting time to treatment, namely in France, Portugal, Canada, US, and Italy. Sixteen indicators (38%) signalled stable waiting times and 8 (19%) a reduction in time to treatment. Twenty-one surveys [58,70,84–86,88,108,109,114,117,132–134,141,147–153], 2 studies using administrative data and surveys [121, 138] and 1 prospective study [117] reported on delays in cancer treatment. From those, eleven articles were international studies. All have reported delays or interruptions on different aspects of cancer treatment, namely in Canada, France, Germany, Italy, Netherlands, US, and UK.

Fifty articles reported on “changes in treatment”, resulting in 304 indicators collated. A total of 106 indicators (19 articles [50,52,65,75,77,90,96–98,101,102,106,118,121,135,137,138,142,154]) were collected from administrative data, 13 indicators from registry data [134, 138] and 185 were survey-base information [13,46,58,70,78,85,88,113–116,126,130,141,145,148–151,155–159]. Changes in treatment are summarized in Fig. 3.

**Fig. 3.**
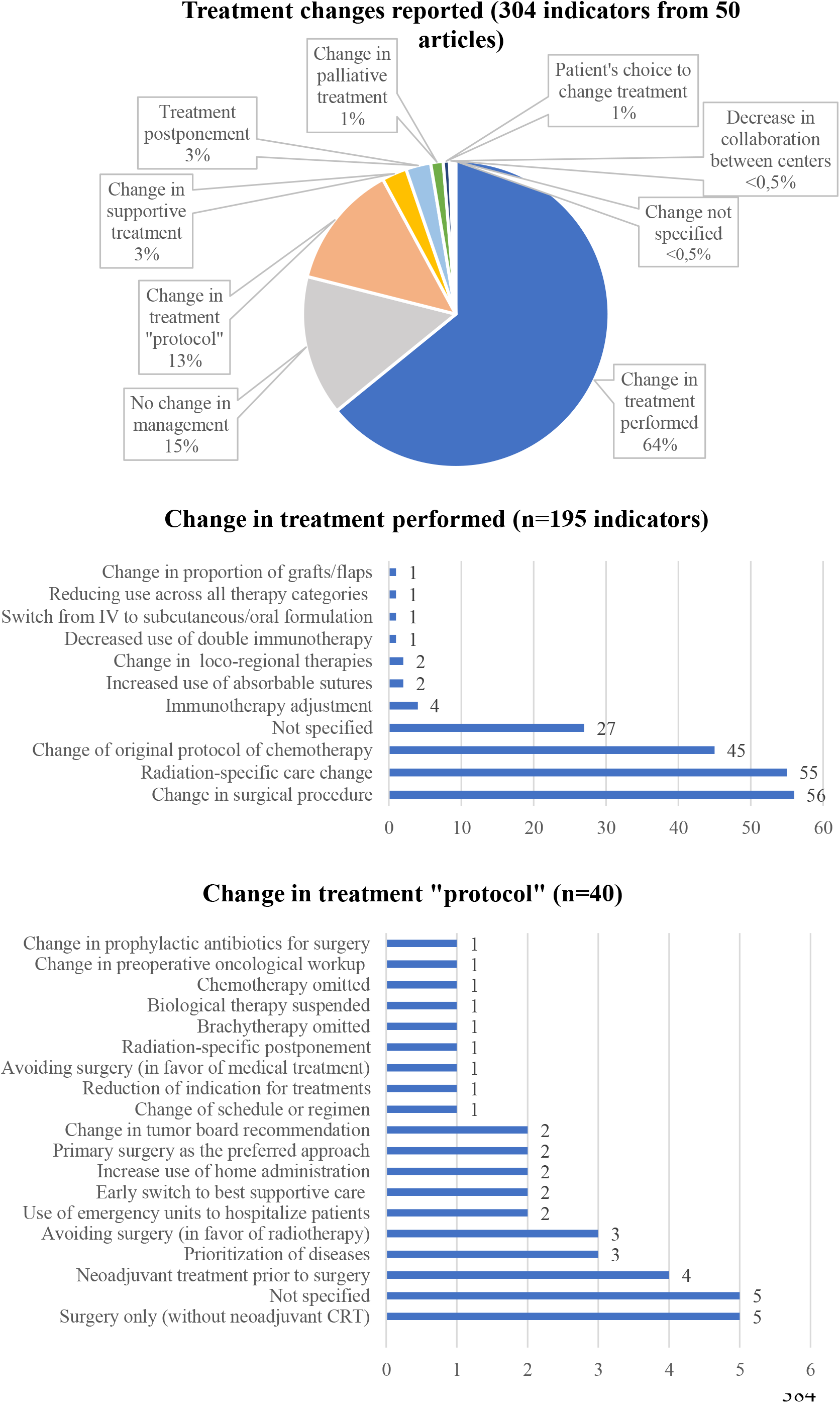
Treatment changes reported in the articles. IV: intravenous; CRT: chemotherapy.

Modifications in treatment were reported in 195 (64%) of indicators. The most frequent changes in treatment were in surgical procedures (n=56 indicators, 29%), radiation-specific changes (n=55 indicators, 28%) and change of original protocol of chemotherapy (n=45 indicators, 23%). Modifications documented in surgical procedures were a decrease in use of laparoscopic surgery together with an increase of open or radical surgery, an increase in stoma formation rate, and a decrease of immediate breast reconstruction rate in breast cancer patients. The radiation-specific care variations identified were radiotherapy hypofractionation, treatment disruptions, increase in short-course treatments, and physicians being less likely to prescribe adjuvant radiotherapy.

Treatment “protocol” changes were reported by 40 indicators. The most frequent change was performing surgery without neoadjuvant chemotherapy (Fig. 3).

A total of 12 indicators from 4 articles [42,55,93,99] reported on the “number of visits and admissions” of cancer patients to the hospital. Of these indicators, 10 (83%) signalled a decreasing trend. Two international surveys [55, 85] to health professionals have also disclosed a decrease in the number of oncology unit hospitalizations.

Regarding the “utilization of telemedicine”, 4 indicators were identified from 4 articles [68,86,103,152]. Fourteen surveys [13,58,70,109,112–115,134,141,145,149,152,157] provided information on telemedicine use, from which ten were international studies. Three were patients’ surveys. All the survey-based information and quantitative indicators reported an increase in the use of telehealth to provide cancer care. One article [160] assessed patient and providers’ satisfaction regarding the use of telemedicine in rehabilitation of cancer patients. The proportion of patients that provided good feedback ranged from 63% to 84%, and the physicians’ perspective was also satisfactory, ranging from 66% to 83% of physicians reporting positive feedback. Four international surveys [115,141,145,150] addressed the implementation of virtual multidisciplinary tumor boards, showing a marked increase in the use of web-based platforms.

Two main outcomes were addressed in the included articles: “procedures and surgical outcome measures” and “mortality”. Fifteen indicators from 8 articles [93,97–103] conveyed information regarding procedures’ outcomes. From these indicators, 11 (73%) showed similar complication rates. One Italian survey [127] has also documented a stable number of complications after esophageal resections.

With respect to mortality in cancer patients, 24 indicators were identified from 4 articles. Twenty of these indicators resulted from one Portuguese study [105], the other 3 indicators showed a stable postoperative death rate in patients with head and neck cancer (France) [102], and a stable in-hospital mortality rate for orthopedic tumors at the traumatology department (Germany) [123]. One Turkish study [100] documented increased mortality in occlusive colorectal cancers patients.

The trends of the indicators are summarized in Fig. 4.

**Fig. 4.**
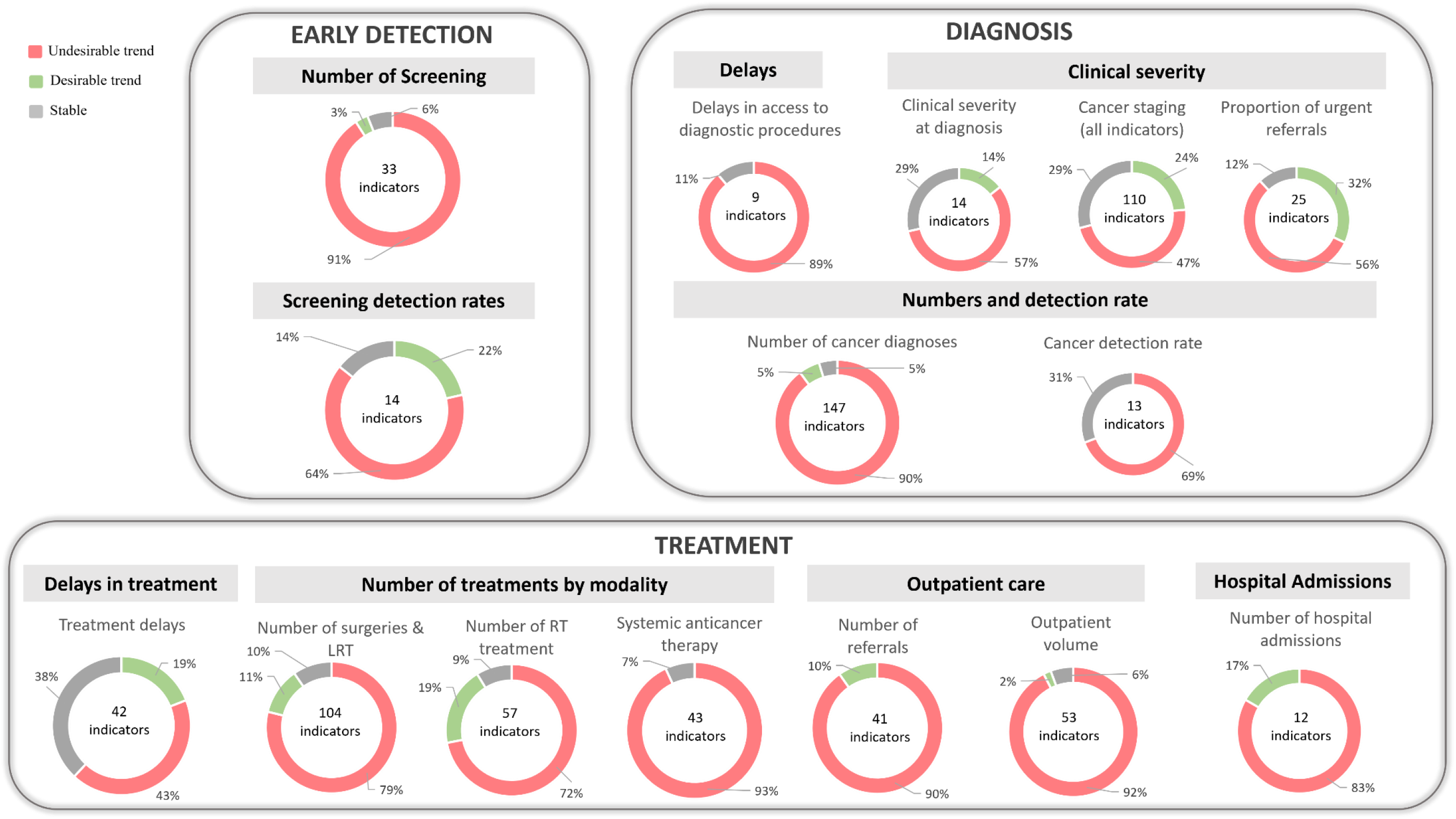
Number of cancer care indicators and trends per category (%), according to the cancer care pathway

## Discussion

In this study, we performed a scoping review to identify the indicators used in the literature to measure the impact of the COVID-19 pandemic on the cancer care pathway from January to June 2020, and the changes in the quality of care signalled by these indicators. We identified a total of 135 articles, with a total of 917 quantitative indicators, reporting on 94 countries. Changes in the quality of care are spread across the care pathway. This performance information suggests capacity constraints, it shows quick adaptations and innovations in cancer management. If collected in near-real-time and processed into actionable information, it would allow monitoring changes in care during the current pandemic and in future events more efficiently, supporting timely and adequate responses.

Our findings signal a major impact on the diagnosis of cancer: a decreasing trend in the number of screenings, diagnostic procedures, and, consequently, in the number of cancer diagnoses, resulting in increasing screening detection rates, and delays in diagnostic care. These result from the reduction of screening programs [4], the cancelation of elective procedures [22], and patients’ avoidance of going to healthcare facilities [161, 162]. Previous studies documented the decreasing number of primary care consultations for a wide range of clinical conditions [163, 164] and a relevant fall in new cancer diagnoses in primary care [76].

Trends regarding cancer staging at diagnosis portray a mixed picture. The reduction in the number of elective diagnostic procedures could explain stage-shifts in cancer presentation. We also found an increasing trend in the proportion of urgent/emergent cancer referrals and in the proportion of emergent procedures, as well as evidence of patients presenting with a more severe clinical condition to the hospital than before the pandemic. These trends suggest patients with advanced-stage cancers continued to seek care. It could also signal those patients waited longer before receiving care, with potential deleterious outcomes. These results highlight the need to closely monitor the impact of the COVID-19 pandemic on shifts in cancer staging at diagnosis and the relevance of collecting this data systematically, which is a reality only in a few OECD countries, such as the Netherlands and Slovenia [165].

With respect to treatment, about 40% of the indicators reporting on waiting times to treatment signalled an increasing trend, we observe a decreasing trend in the volume of the three modalities of cancer treatment, and a large number and diversity of information reporting on treatment changes. These results reveal the remarkable influence of the pandemic on patterns of cancer care in the first half of 2020. These changes result from postponements of care decided by physicians to decrease patients’ exposure to hospitals [113], switch to audio- or video-consultations, and deferrals and treatment modifications guided by updated recommendations by many medical societies [11, 12]. Almost two-thirds of the indicators reported changes in treatment, which shows that providers have quickly adapted their care practices, which strengthens the argument for the need for monitoring closely these changes. While some of these care modifications could be learning opportunities for the future, this information should be standardised, transparent, and timely, allowing to appraise the modifications in care provided during crises regarding access, quality, and outcomes.

The increase in telemedicine utilization we report is a generalized trend across a range of medical specialties [4] and it constitutes a hallmark of the innovation triggered by this pandemic. Albeit the positive feedback by physicians and patients we report, telehealth risks to increase inequalities in access to care [166, 167].

Short-term oncological outcomes were addressed by a few indicators and are reported as being stable, which is in line with a recent international cohort study including 61 countries and 15 tumor types [22]. However, deferred care will most likely lead to worse long-term outcomes, which needs to be monitored. Attempts to quantify this impact were developed, for instance, by a British nation-wide modelling study where the authors estimated a total of 59 204–63 229 additional years of life lost attributed to four major cancers [168], compared with pre-pandemic data.

Previous works addressed the impact of the COVID-19 pandemic on regional or national settings [5, 50], on specific cancers [5,6,169], specific stages of the care pathway [54], or treatment modalities [22]. This scoping review provides a summary of cancer care performance indicators, concerning various diseases, from early detection to the treatment phase of the care pathway, within OECD countries. Additionally, we report changes in the quality of cancer care based on indicators’ trends, from January to June 2020, which constitutes an innovative approach to assess changes in healthcare performance.

Our study has some limitations. The heterogeneity of study designs, populations, diseases, indicators, and indicators’ definitions do not allow application of a meta-analysis approach and quantifying the real impact of the pandemic on cancer care. We also recognize potential bias that could arise from the inclusion of survey data, which rely on what respondents recall and report. However, a relevant number of surveys were conducted by international societies and networks of providers, which allows for the collection of credible information regarding changes in care. Surveys also show how medical societies and countries were unable to obtain these data using current health information systems and data structures. We acknowledge the patients’ perspective is not comprehensive since only a few surveys included patient-reported outcomes and experiences of care measures. The cancer care pathway is not complete in this study, as it did not address primary prevention, rehabilitation, and palliative care. Nonetheless, we provide a thorough overview of the cancer care pathway, from diagnosis to outcomes.

The distribution of studies per country is not homogenous in this study, which does not allow to generalize these trends to all included countries or to fully assess disparities in cancer care between OECD countries. Care inequalities could have been exacerbated during this pandemic, which needs to be further studied regarding cancer care.

The COVID-19 pandemic keeps evolving until the present time and postponing of care was reported in some countries by the end of 2021 and the beginning of 2022 [170, 171], which means that this impact is adding up. Figures concerning new COVID-19 cases, mortality, and vaccination coverage, are presented daily to the public since the beginning of 2020. Additional and considerable efforts are needed to expose the effects caused by this pandemic in non-COVID-19 care, and some of the indicators we present could be useful to convey that message.

As new waves keep evolving, it is crucial to monitor performance indicators, such as shifts in cancer staging or worsening of outcomes, closely and promptly. Also, the link between structure, process, and (short- and long-term) outcome indicators should be undertaken to allow an accurate and timely evaluation of the changes in the care provided during crises (and in regular times). Tools like the “Time to Act Data Navigator” [172] developed by the European Cancer Organization and the “Global Cancer Observatory” [173] signal the ambition to address this lack of standardised and regular collection of data and indicators. Within the scope of Europe’s Beating Cancer Plan, the European Cancer Inequalities Registry [20] aims to monitor inequalities across Europe, by providing reliable data on cancer prevention and care.

## Conclusion

Our results provide a summary of performance indicators used in the literature to assess the cancer care pathway from January 2020 to June 2020, and the changes in the quality of care signalled by these indicators. This study shows that health systems struggled to ensure the continuity in the quality of care to cancer patients in OECD countries. This information could inform on the bottlenecks of the cancer care pathway and contribute to identifying disparities between and within countries, as well as moments for intervention during the evolving pandemic and in future crises. Further research and investment are necessary to generate system-wide oriented intelligence and strengthen data infrastructures worldwide, to support timely and adequate health policy responses.

## Supporting information

Supplemental File S2 Doc

## Data Availability

The data underlying this article are available in Zenodo.org, at https://doi.org/10.5281/zenodo.6129839.

## Acknowledgements

The authors wish to thank Wichor Bramer from the Erasmus MC Medical Library for developing and updating the search strategies used in this study.

## Ethics approval and consent to participate

Not required.

## Authors’ contributions

All authors contributed to conceptualize the study. ASC and MdL performed the data collection. ASC performed the data analysis and drafted the article. All authors provided feedback and contributed to revising the manuscript. All authors approved the final version of the manuscript.

## Supporting information

**S1 Table – Preferred Reporting Items for Systematic reviews and Meta-Analyses extension for Scoping Reviews (PRISMA-ScR) Checklist**

**S2 Doc – Search strategy**

**S3 Table – Data Extraction Form**

**S4 Table – Characteristics of the studies included in the review, and from which indicators were extracted and collated**

