## Supplemental File S2 Doc for "Changes in the quality of cancer care as assessed through performance indicators during the first wave of the COVID-19 pandemic in 2020: a Scoping Review"

**S2 Doc – Search strategy**

| **Database searched** | **via** | **Years of coverage** | **Records** | **Records after duplicates removed** |
| --- | --- | --- | --- | --- |
| Embase | Embase.com | 1971 - Present | 4282 | 4206 |
| Medline ALL | Ovid | 1946 - Present | 4429 | 2071 |
| **Total** | | | **8711** | **6277** |

**Embase.com	4282**

(pandemic/mj/exp OR 'natural disaster'/mj/exp OR disaster/mj/exp OR hurricane/mj/exp OR earthquake/mj/de OR 'coronavirus disease 2019'/mj/de OR 'Severe acute respiratory syndrome coronavirus 2'/mj/de OR (pandemic* OR disaster* OR hurricane* OR flood OR flooding OR earthquake* OR coronavirus-disease-2019 OR covid19 OR covid-19 OR sars-cov-2 OR 2019-novel-coronavirus OR 2019-ncov OR lockdown OR lock-down OR typhoon* OR (natural NEAR/3 hazard*)):ti) AND ('non communicable disease'/de OR neoplasm/exp OR 'malignant neoplasm'/de OR 'cancer patient'/exp OR 'heart disease'/exp OR cardiology/de OR 'neurologic disease'/de OR 'cancer screening'/de OR 'cancer surgery'/de OR (('primary medical care'/exp OR 'general practitioner'/de OR 'general practice'/de OR 'family medicine'/de OR 'emergency ward'/exp OR 'emergency care'/de) AND ('health care utilization'/exp OR consultation/de)) OR 'diabetes mellitus'/exp OR 'rheumatic disease'/exp OR 'mental health service'/de OR 'cardiovascular disease'/exp OR 'cerebrovascular disease'/exp OR 'chronic kidney failure'/exp OR 'obstructive airway disease'/exp OR 'chronic respiratory tract disease'/de OR 'cerebrovascular accident'/exp OR hypertension/exp OR 'chronic disease'/de OR 'neurologic disease'/exp OR (((non-communicab* OR noncommunicab*) NEAR/3 disease) OR cancer OR ((heart OR cardiovascul* OR cerebrovascul*) NEAR/3 (disease* OR patient* OR emergenc*)) OR cardiolog* OR neurolog* OR oncolog* OR ((((emergency OR acute) NEAR/3 (ward* OR care OR department*)) OR (general NEXT/1 (practi*)) OR ((primary OR family) NEXT/2 (healthcare* OR care OR doctor*))) AND (utilization* OR utilisation* OR delay* OR time OR visit* OR consultation* OR barrier* OR access* OR challenge* OR vulnerab* OR attendan* OR hesita*)) OR diabet* OR rheumat* OR (mental NEAR/3 (health-care* OR health-service* OR healthcare*)) OR (chronic NEAR/3 (kidney OR renal OR respirator* OR lung* OR pulmonar*) NEAR/3 (failure OR disease*)) OR asthma* OR bronchitis OR ((lung* OR pulmonar*) NEAR/3 (emphysem*)) OR ((cerebrovascul* OR cerebro-vascul*) NEAR/3 accident*) OR stroke OR hypertens* OR chronic-disease* OR (surg* NEAR/3 emergenc*)):ab,ti) AND ('disease exacerbation'/mj/de OR 'recurrent disease'/mj/de OR 'avoidance behavior'/mj/de OR 'recurrence risk'/mj/de OR 'undiagnosed disease'/mj/de OR 'diagnostic error'/mj/de OR 'diagnostic delay'/mj/de OR 'delayed diagnosis'/mj/de OR 'therapy delay'/mj/de OR 'health care utilization'/mj/de OR mortality/mj/exp OR morbidity/mj/exp OR 'health care quality'/mj/exp OR screening/mj OR 'mass screening'/exp/mj OR 'screening test'/mj OR 'palliative therapy'/exp/mj OR 'health care delivery'/mj OR 'health care access'/exp/mj OR 'surgical volume'/mj OR rehabilitation/exp/mj OR 'performance indicator'/de OR (impact* OR influence* OR affect* OR effect* OR exacerbate* OR (Disease NEAR/3 progression*) OR recur* OR avoid* OR postpon* OR post-pon* OR implication* OR ((diagnos* OR therap* OR treatment* OR hospitali* OR services OR visit*) NEAR/3 (error* OR delay* OR fewer* OR drop OR decline* OR continuit* OR decrease* OR increase* OR reduc*)) OR ((health-care OR healthcare) NEAR/3 (utilizat* OR utilisat* OR use)) OR mortalit* OR morbidit* OR ((health-care OR healthcare) NEAR/3 (quality)) OR screening OR (service* NEAR/3 ( disrupt* OR continu*)) OR palliat* OR unmet OR rehabilitation*):ti OR (((performance* OR outcome*) NEAR/3 (indicator*)) OR ((health-care OR healthcare) NEAR/3 (access* OR deliver* OR output*)) OR ((surgical OR surgeries OR procedures) NEAR/3 (volum* OR number*))):ab,ti) NOT (model/exp/mj OR (model*):ti) NOT ([conference abstract]/lim AND [2000-2019]/py) NOT ([animals]/lim NOT [humans]/lim) NOT ('health care personnel'/exp/mj OR pregnancy/exp/mj OR 'pregnant woman'/mj OR (((COVID-19 OR COVID19 OR coronavirus* OR corona-virus* OR SARS-CoV-2) NEAR/3 (outcome* OR case* OR patient* OR pneumoni* OR progression* OR prognos* OR mortalit* OR fatal* OR sever* OR vaccine*)) OR pregnan* OR ((healthcare OR health-care OR medical) NEAR/3 (personnel* OR staff OR worker* OR workforce* OR work-force*)) OR doctor* OR nurse* OR physician*):ti OR ((COVID-19 OR COVID19 OR coronavirus* OR corona-virus* OR SARS-CoV-2) NEAR/3 (mortalit* OR outcome* OR fatal*)):ab) NOT ((child/exp NOT adult/exp) OR (pediatr* OR paediatr* OR child* OR infan* OR adolescen*):ti) NOT ('practice guideline'/de OR (guideline*):ti)

**Medline ALL Ovid	4429**

(*Pandemics/ OR *Natural Disasters/ OR *Cyclonic Storms/ OR *Earthquakes/ OR *COVID-19/ OR *SARS-CoV-2/ OR (pandemic* OR disaster* OR hurricane* OR flood OR flooding OR earthquake* OR coronavirus-disease-2019 OR covid19 OR covid-19 OR sars-cov-2 OR 2019-novel-coronavirus OR 2019-ncov OR lockdown OR lock-down OR typhoon* OR (natural ADJ3 hazard*)).ti.) AND (Noncommunicable Diseases/ OR exp Neoplasms/ OR exp Heart Diseases/ OR Cardiology/ OR exp Nervous System Diseases/ OR Early Detection of Cancer/ OR exp Diabetes Mellitus/ OR exp Rheumatic Diseases/ OR exp Mental Health Services/ OR exp Cardiovascular Diseases/ OR exp Cerebrovascular Disorders/ OR exp Kidney Failure, Chronic/ OR Lung Diseases, Obstructive/ OR exp Stroke/ OR exp Hypertension/ OR Chronic Disease/ OR ((exp Emergency Medical Services/ OR exp Emergency Treatment/ OR General Practitioners/ OR General Practice/ OR Family Practice/) AND (Patient Acceptance of Health Care/ OR "Referral and Consultation"/)) OR (((non-communicab* OR noncommunicab*) ADJ3 disease) OR cancer OR ((heart OR cardiovascul* OR cerebrovascul*) ADJ3 (disease* OR patient* OR emergenc*)) OR cardiolog* OR neurolog* OR oncolog* OR ((((emergency OR acute) ADJ3 (ward* OR care OR department*)) OR (general ADJ (practi*)) OR ((primary OR family) ADJ2 (healthcare* OR care OR doctor*))) AND (utilization* OR utilisation* OR delay* OR time OR visit* OR consultation* OR barrier* OR access* OR challenge* OR vulnerab* OR attendan* OR hesita*)) OR diabet* OR rheumat* OR (mental ADJ3 (health-care* OR health-service* OR healthcare*)) OR (chronic ADJ3 (kidney OR renal OR respirator* OR lung* OR pulmonar*) ADJ3 (failure OR disease*)) OR asthma* OR bronchitis OR ((lung* OR pulmonar*) ADJ3 (emphysem*)) OR ((cerebrovascul* OR cerebro-vascul*) ADJ3 accident*) OR stroke OR hypertens* OR chronic-disease* OR (surg* ADJ3 emergenc*)).ab,ti.) AND (*Disease Progression/ OR exp * Recurrence/ OR *Undiagnosed Diseases/ OR exp *Diagnostic Errors/ OR *Delayed Diagnosis/ OR * Time-to-Treatment/ OR *Patient Acceptance of Health Care/ OR exp *Mortality/ OR exp *Morbidity/ OR exp *Quality of Health Care/ OR *Early Detection of Cancer/ OR *Mass Screening/ OR *Palliative Care/ OR exp *Delivery of Health Care/ OR *Health Services Accessibility/ OR *Rehabilitation/ OR (impact* OR influence* OR affect* OR effect* OR exacerbate* OR (Disease ADJ3 progression*) OR recur* OR avoid* OR postpon* OR post-pon* OR implication* OR ((diagnos* OR therap* OR treatment* OR hospitali* OR services OR visit*) ADJ3 (error* OR delay* OR fewer* OR drop OR decline* OR continuit* OR decrease* OR increase* OR reduc*)) OR ((health-care OR healthcare) ADJ3 (utilizat* OR utilisat* OR "use")) OR mortalit* OR morbidit* OR ((health-care OR healthcare) ADJ3 (quality)) OR screening OR (service* ADJ3 ( disrupt* OR continu*)) OR palliat* OR unmet OR rehabilitation*).ti. OR (((performance* OR outcome*) ADJ3 (indicator*)) OR ((health-care OR healthcare) ADJ3 (access* OR deliver* OR output*)) OR ((surgical OR surgeries OR procedures) ADJ3 (volum* OR number*))).ab,ti.) NOT (exp * Models, Theoretical/ OR model*:ti) NOT (exp animals/ NOT humans/) NOT (exp * Health Personnel/ OR exp * Attitude of Health Personnel / OR exp * pregnancy/ OR * pregnant woman/ OR (((COVID-19 OR COVID19 OR coronavirus* OR corona-virus* OR SARS-CoV-2) ADJ3 (outcome* OR case* OR patient* OR pneumoni* OR progression* OR prognos* OR mortalit* OR fatal* OR sever* OR vaccine*)) OR pregnan* OR ((healthcare OR health-care OR medical) ADJ3 (personnel* OR staff OR worker* OR workforce* OR work-force*)) OR doctor* OR nurse* OR physician*).ti. OR ((COVID-19 OR COVID19 OR coronavirus* OR corona-virus* OR SARS-CoV-2) ADJ3 (mortalit* OR outcome* OR fatal*)).ab.) NOT (((exp child/ OR exp infant/) NOT exp adult/) OR (pediatr* OR paediatr* OR child* OR infan* OR adolescen*).ti.) NOT (Practice Guideline/ OR Practice Guidelines as Topic/ OR (guideline*).ti.)
