## Supplemental File S3 Table for "Changes in the quality of cancer care as assessed through performance indicators during the first wave of the COVID-19 pandemic in 2020: a Scoping Review"

**Data Extraction Form**

**Form 1: General study characteristics**

| Article ID | Number identifying each paper |
| --- | --- |
| Article source | Systematic search |
|  | Reference mining |
| Clinical area |  |
| Disease/care delivered |  |
| Country |  |
| Study design | Randomized controlled trial |
|  | Prospective cohort |
|  | Retrospective cohort |
|  | Case-control |
|  | Cross-sectional |
|  | Case report |
|  | Systematic review |
|  | Meta-analysis |
|  | Scoping review |
|  | Survey |
| Peer-reviewed | Yes |
|  | No |
| Excluded: (with reason for exclusion) | Yes (reason) / No |
| Time period studied |  |
| Comparison period | 2020 vs. 2019 |
|  | 2020: pre- versus post-COVID |
|  | 2020 vs. previous years |
| Article title |  |
| Year of publication |  |
| Journal |  |
| Authors |  |
| Paper URL |  |
| Language |  |
| Abstract |  |

**Form 2: Indicator related information**

| Paper ID | Number identifying each paper |
| --- | --- |
| Clinical area |  |
| Disease/care delivered |  |
| Country |  |
| Data source | Population-level data |
|  | Adverse incident data |
|  | Clinical data |
|  | Costing data |
|  | Claims data |
|  | Survey data |
|  | Registry data |
|  | Prescribing data |
|  | Claims data |
| Database |  |
| Sample size | Number of centers |
|  | Number of patients/procedures/responses/events |
| Indicator title |  |
| Self-reported | Yes |
|  | No |
| Numerator (inclusion criteria) |  |
| Numerator (exclusion criteria) |  |
| Denominator |  |
| Indicator results from statistical model | Yes |
|  | No |
| Periods being compared | 2020 vs. 2019 |
|  | 2020: pre- versus post-COVID |
|  | 2020 vs. previous years |
| Trend reported by indicator | Increase |
|  | Stable |
|  | Decrease |
| Numerical value before pandemic |  |
| Numerical value during pandemic |  |
| Units relating to numerical value | (eg: ng/L, days, admissions) |
| Magnitude (%) | *Computed as percent change:*  ((value during pandemic - value before pandemic) /  (value before pandemic)) * 100 |
| Main results (if reported) | Relative risk ratio |
|  | Odds ratio |
|  | Incidence rate ratio |
| Other results reported (free field) |  |
| Delivery of care pathway | Access / admission |
|  | Diagnostic |
|  | Treatment |
|  | Follow-up / Outpatient care |
|  | Other |
|  | Outcomes |
| Dimensions of performance | Access |
|  | Quality – effectiveness |
|  | Quality – safety |
|  | Quality – patient-centredness |
| Donabedian's framework: | Structure |
|  | Process |
|  | Outcome |
